# Beyond standardized health-related quality of life measures: Additional symptoms and problems reported through the EORTC WISP instrument in the EUonQoL-Kit study across European cancer populations

**DOI:** 10.64898/2026.11.23.26368179

**Authors:** Leslye Rojas-Concha, Mathilde Adsersen, Giovanni Apolone, Anne Brédart, Federico Borgogni, Cinzia Brunelli, Augusto Caraceni, Ana Carneiro, Luana Caselli, Tudor Ciuleanu, Michal Chovanec, Carina Dantas, Philip Robert Debruyne, Daniela Doege, Montserrat Ferrer, Theres Fey, Olatz Garin, Alexandra Gilbert, Marianne Gronlie Guren, Hayat Hamzeh, Maiken Bang Hansen, Orsolya Horváth, Sara Iadecola, Angelos Kassianos, Tomáš Kazda, Ekaterina Kldiashvili, Sandra Lejniece, Iwona Ługowska, Chiara Marzorati, Johanna Mattson, Inke Minnée-van Braak, Milana Mitrić Asković, Emma Nicklin, Helidon Nina, Helle Pappot, Morten Aagaard Petersen, Ivaylo Petrov, Vadim Pogonet, Gabriella Pravettoni, Ivica Ratoša, Alessandro Rizzo, Marie Robert, Gerald Sendlhofer, Antonio Tanzilli, Hugo Vachon, Vahur Valvere, Ines Vaz-Luis, Mogens Groenvold EUonQoL consortium

**Author notes:** **Corresponding author**: Leslye Rojas-Concha, Palliative Care Research Unit, Department of Geriatric and Palliative Medicine GP, Copenhagen University Hospital - Bispebjerg and Frederiksberg. Bispebjerg Bakke 23, DK-2400 Copenhagen NV, Denmark.

## Abstract

**Background:** Standardized health-related quality of life (HRQoL) measures may not capture all symptoms/problems relevant to people with cancer. The European Oncology Quality of Life toolkit (EUonQoL-Kit), co-created with multiple stakeholders to inform health policy across Europe, combines standardized closed-ended patient-reported outcome (PRO) items with the European Organisation for Research and Treatment of Cancer (EORTC) open-ended Write In three Symptoms/Problems (WISP) instrument. We aimed to identify symptoms/problems reported via WISP beyond the EUonQoL-Kit, examine differences in background characteristics between WISP respondents and non-respondents, and compare additional symptom/problem reporting across three target cancer populations: patients in active treatment, survivors, and patients in palliative care.

**Methodology:** This descriptive analysis used data from adults with current or previous cancer diagnoses participating in a multicenter validation study across 32 European countries (22 August 2024–7 July 2025). WISP responses were categorized, and subsequently their prevalence and severity were calculated. Participant characteristics and the ten most frequently reported additional symptoms/problems were compared across groups using t-tests, chi-square tests, or Fisher’s exact tests, as appropriate.

**Results:** Among 4,284 included participants, 519 (12.1%) reported 954 symptoms/problems on WISP, of which 383 (40.1%) were not covered by EUonQoL-Kit v2 closed-ended items. Women, participants from Western and Northern Europe, highly educated and those diagnosed with breast cancer were more likely to respond to the WISP than other subgroups (all p <0.05). No statistically significant differences were found in the prevalence of common additional symptoms/problems (p≥0.130). Descriptively, numbness/tingling, skin problems and bleeding were most reported by patients in active treatment; indigestion, incontinence, and vision problems in survivors; and edema, cough and itching in palliative care.

**Conclusions:** The EORTC WISP complemented standardized PRO assessment by capturing additional symptoms/problems. Integrating open-ended WISP reporting into the final toolkit may enhance HRQoL assessment across European cancer populations while identifying population-specific concerns without expanding predefined items for all respondents.

## Introduction

There is growing interest in the use of Patient-Reported Outcome (PRO) instruments to assess symptoms and Health-Related Quality of Life (HRQoL), as they provide direct assessment of patients’ health status from their own perspective [1, 2]. The use of PRO assessment at individual level has shown to improve cancer care by enabling earlier detection and management of symptoms, enhancing patient-clinician communication, and increasing patient satisfaction [3, 4]. At a healthcare system level, aggregated PRO data can be used to assess the effectiveness of healthcare services and programs, thereby facilitating evidence-based policy decision-making [5]. At the same time, PRO assessment face challenges, including potential response bias, methodological flaws, and workflow burdens. For example, patients may underreport symptoms to avoid treatment modification or may overreport symptoms to please clinicians [6]. These limitations highlight the importance of developing comprehensive, patient-centered approaches to PRO assessment. In this context, the EU-funded project “Quality of Life in Oncology: measuring what matters to cancer patients and survivors in Europe (EUonQoL)” developed the EUonQoL-Kit, a unified, patient-centered toolkit for assessing HRQoL in cancer patients and survivors across Europe to support health policy decision-making [7].

The EUonQoL-Kit was co-created with patients as co-researchers, using a mixed-method approach. The development is described in detail elsewhere and included systematic reviews [8–11] qualitative patient interviews [12], a Delphi consensus study [13] and usability testing [14]. Findings from these studies were triangulated and reviewed with key stakeholders during several international, multidisciplinary consensus meetings, selecting the most relevant HRQoL subdomains and items to form the final EUonQoL-Kit v2 to be tested in the validation study [14]. The decisions for inclusion and exclusion of subdomains and items focused on the overall aim of the EUonQoL-Kit and its use in health policy decision-making. The EUonQoL-Kit was translated and culturally adapted into the 31 target European languages [15].

The EUonQoL-Kit v2 tested in the validation study consisted of six questionnaires, including a dynamic version using Computer Adapted Testing (CAT) and a static version for each of the three target cancer populations: patients in active treatment (i.e., receiving or recently completed antineoplastic treatment) survivors and patients in palliative care [14]. Both versions also included the open-ended ‘Write In three Symptoms/Problems’ (WISP) instrument, developed by the European Organisation for Research and Treatment of Cancer (EORTC). The WISP was designed to give patients the opportunity to report up to three additional symptoms or problems that were not captured by the preceding questionnaire [16]. A final open-ended question on overall quality of life was included in all questionnaires (Having completed the questionnaire, what do you feel most impacts your Quality of Life?) [14].

Although comprehensive HRQoL measures can cover several symptoms/problems, they may not capture concerns that are uncommon, disease- or treatment-specific, or particularly relevant to people with cancer. The WISP provides a simple way for patients to bring these additional concerns to light, without requiring all patients to complete a longer list of questions that may not be important to them. Previous research has shown that WISP can identify symptoms/problems that are not covered by standardized cancer questionnaires [17, 18].

In the EUonQoL-Kit v2 validation study, the EORTC WISP instrument was included to explore whether patients would report additional symptoms/problems beyond those captured by the toolkit. Thus, the objectives of this study were to (1) identify symptoms/problems reported on the WISP in addition to those covered by the EUonQoL-Kit v2, (2) assess whether participants who reported symptoms/problems on the WISP differed from those who did not in terms of their background characteristics, and (3) explore differences in the reporting of these additional symptoms/problems among the three target populations.

## Methods

### Study design and target populations

The EUonQoL-Kit v2 was tested in a multicenter observational cross-sectional study. This validation study, was carried out between 22 August 2024 and 07 July 2025 in 45 cancer centers from 25 EU Member States and 7 associated countries [15]. The validation study aimed to recruit 4,500 cancer patients and survivors across participating countries. To assess test-retest reliability of the EUonQoL-Kit v2, a randomly selected 10% subsample of the overall study population, stratified by target populations, was invited to complete the instrument a second time at least an hour after the initial assessment [15].

The EUonQoL-Kit validation study included individuals aged 18 years or older who had a current or past cancer diagnosis and belonged to one of the three target populations: a) active treatment (patients receiving or recently completed curative treatment for early-stage cancer or non-curative, disease-controlling or life-prolonging treatment for advanced/metastatic cancer), b) survivors (disease-free patients without evidence of active cancer and at least one year off active treatment), and c) palliative care (patients with advanced cancer with limited prognosis <12 months, specialist palliative care referral, or treatment aimed solely at symptom control). Participants were required to be native or fluent speakers of the target language and to provide written informed consent. Individuals were considered ineligible to participate if, in the judgment of the local investigator responsible for patient screening, cognitive impairment prevented completion of the questionnaire [15].

### The EUonQoL-Kit and the EORTC WISP instrument

Both dynamic and static versions of the EUonQoL-Kit v2 were electronically administered. The dynamic version incorporated the EORTC CAT Core, which is based on Item Response Theory (IRT) [19], whereas the static version comprised questionnaires with traditional pre-selected set of items [15].

The content of the EUonQoL-Kit v2 was structured according to a five-domain conceptual framework encompassing physical health, psychological well-being, social health, overall health, and health care [14]. The subdomains included in the EUonQoL-Kit v2, together with the number of items included for each of the three target populations are presented in Table 1. While the overall conceptual framework remained similar across populations, some variations in content were introduced to reflect population-specific needs. In particular, the palliative care questionnaires included fewer questions addressing psychical functioning, fear of recurrence, family & relationships, financial difficulties, the impact of cancer on life role, and did not include the question on lifestyle changes and body image (Table 1) [14].

**Table 1.** Content and number of items in the dynamic and static versions of the EUonQoL-Kit v2.

|  | EUonQoL-Kit v2 |  |  |  |  |  |
|---|---|---|---|---|---|---|
| **Subdimensions** | **Dynamic version** |  |  | **Static version** |  |  |
|  | Active treatment | Survivors | Palliative care | Active treatment | Survivors | Palliative care |
| ***Physical health*** |  |  |  |  |  |  |
| Mobility & activity/ Physical functioning | 7 | 7 | 7 | 3 | 3 | 2 |
| Pain | 6 | 6 | 6 | 2 | 2 | 2 |
| Energy/Fatigue | 6 | 6 | 6 | 2 | 2 | 2 |
| Sleeping problems | 6 | 6 | 6 | 1 | 1 | 1 |
| Lack of appetite | 1 | 1 | 1 | 1 | 1 | 1 |
| Nausea/Vomiting | 1 | 1 | 1 | 1 | 1 | 1 |
| Constipation | 1 | 1 | 1 | 1 | 1 | 1 |
| Diarrhea | 1 | 1 | 1 | 1 | 1 | 1 |
| Dyspnea | 1 | 1 | 1 | 1 | 1 | 1 |
| Sexual life/intimacy | 1 | 1 | 1 | 1 | 1 | 1 |
| Body image |  | 1 |  |  | 1 |  |
| Symptom worries | 3 | 3 | 3 | 3 | 3 | 3 |
| Symptom side effects | 1 | 1 | 1 | 1 | 1 | 1 |
| ***Psychological wellbeing*** |  |  |  |  |  |  |
| Anxiety & worry/ Emotional functioning | 7 | 7 | 7 | 3 | 3 | 3 |
| Spirituality | 1 | 1 | 1 | 1 | 1 | 1 |
| Future outlook | 1 | 1 | 1 | 1 | 1 | 1 |
| Cognitive functioning | 1 | 1 | 1 | 1 | 1 | 1 |
| Fear of recurrence | 2 | 2 | 1 | 2 | 2 | 1 |
| ***Social health*** |  |  |  |  |  |  |
| Family & relationships | 2 | 2 | 1 | 2 | 2 | 1 |
| Social functioning | 6 | 6 | 6 | 3 | 3 | 3 |
| Social role & activities/ Role functioning | 6 | 6 | 6 | 2 | 2 | 2 |
| Financial difficulties | 6 | 6 | 6 | 2 | 2 | 1 |
| Impact of cancer on life role | 2 | 2 | 1 | 2 | 2 | 1 |
| Maintenance of independence | 1 | 1 | 1 | 1 | 1 | 1 |
| ***Overall health*** |  |  |  |  |  |  |
| Global health and quality of life | 2 | 2 | 2 | 2 | 2 | 2 |
| Health behavior change | 1 | 1 |  | 1 | 1 |  |
| Open-ended WISP instrument | 1 | 1 | 1 | 1 | 1 | 1 |
| ***Health care*** |  |  |  |  |  |  |
| Care and communication | 5 | 4 | 5 | 5 | 4 | 5 |
| Impact of care pathway | 2 | 2 | 2 | 2 | 2 | 2 |
| Open-ended question on overall quality of life | 1 | 1 | 1 | 1 | 1 | 1 |
| **Total number of items** | **82** | **82** | **78** | **50** | **50** | **44** |

The WISP instrument was administered immediately before the health care domain, which assessed patients’ experiences of health care. It allowed participants to report up to three symptoms/problems that were not covered by the EUonQoL-Kit v2 and to rate their severity on a four-point scale ranging from 1 (“not at all”) to 4 (“very much”)[16].

### Data analysis

The descriptive analysis reported in this study focused exclusively on the WISP results; findings from other EUonQoL-Kit investigations will be reported in future publications.

#### Qualitative analyses

WISP responses were translated from the native official language of each participating country into English using DeepL translator [20] and subsequently reviewed with ChatGPT-4. The translations were then checked by native speakers fluent in English at the respective recruitment sites (except in English-speaking countries) to ensure that participants’ original meanings were accurately preserved, particularly for misspellings, slang, colloquial, or culturally specific expressions not identified by Artificial Intelligence (AI) tools. Any discrepancies were checked against the original responses and corrected during the review. Thus, AI tools supported the translation process, while final verification was performed by the research team.

The final translated WISP responses were codified using the established WISP coding system, which includes a list of 63 symptom/problem categories [21]. Additional codes were created if a symptom/problem did not match any existing category. WISP responses were initially coded by one researcher (LRC) and subsequently reviewed by a second researcher (HH). Any disagreements regarding the assigned codes were identified during the review and discussed within the research team, with final coding decisions reached by consensus.

#### Quantitative analyses

Participant sociodemographic and clinical characteristics were summarized using proportions for categorical variables and means and standard deviations for continuous variables. Characteristics were compared between participants who reported symptoms/problems using the WISP instrument and those who did not, using chi-square test and unpaired t-test, as appropriate. For categorical variables with more than two categories, statistically significant overall group differences were further explored using unadjusted standardized residuals, with |z|> 1.96 indicating subcategories that deviate from the expected frequencies, i.e., contribute particularly to the significant overall difference.

WISP symptom/problem categories were classified according to whether they were covered or not covered by the EUonQoL-Kit. Only WISP responses with a score of 2 (“a little”) or higher were included in the analyses. The prevalence of each symptom/problem category was calculated for the overall population and separately for the three target populations. Symptom/problem severity was categorized according to the WISP response options: ‘a little’ was classified as mild, ‘quite a bit’ as moderate, and ‘very much’ as severe. The proportion of responses falling into each severity category was then calculated across all participants.

We also calculated the mean and median number of symptoms/problems listed on the WISP instrument for all participants completing the EUonQoL-Kit and for those reporting on the WISP instrument, as well as according to the target populations.

The prevalence of the 10 most common symptoms/problems reported on WISP (not covered by the EUonQoL-Kit) were compared among the target populations using Fisher’s exact tests. All statistical tests were performed using a significance level of 0.05. Analyses were conducted using the statistical software Statistical Package for the Social Sciences (SPSS) version 29.

## Results

### Study population

In total 4,302 participants were enrolled in the validation study, of which 4,284 completed the EUonQoL-Kit and were included. Among these participants, 519 (12.1%) reported at least one symptom or problem on the EORTC WISP instrument during the first assessment with the EUonQoL-Kit v2. Of these 519 participants answering the WISP, 193 (37.2%) were patients in active treatment, 172 (33.1%) were survivors and 154 (29.7%) were patients in palliative care

Participant characteristics differed significantly between those who reported symptoms/problems on the WISP and those who did not (Table 2). Chi-square tests showed that women significantly more often reported on WISP than men (13.3% vs. 10.9%). Follow-up post-hoc analyses showed that participants from Western (15.6%) and Northern (14.8%) Europe, those completing post-secondary education (15.9%), and those diagnosed with breast cancer (15.4%) were more likely to respond to the WISP than other participants.

**Table 2.** Participant characteristics who reported symptoms/problems using the WISP instrument compared with those who did not.

| Characteristics | Participants reporting symptoms/problems on WISP |  |  |  |  |  |
|---|---|---|---|---|---|---|
|  |  | Yes (n=519) |  | No (n=3,765) |  |  |
|  | Total | N | % | N | % | *p*-value |
| Patient group |  |  |  |  |  | 0.088 |
| In active treatment | 1,747 | 193 | 11.0 | 1,554 | 89.0 |  |
| Survivors | 1,255 | 172 | 13.7 | 1,083 | 86.3 |  |
| In palliative care | 1,282 | 154 | 12.0 | 1,128 | 88.0 |  |
| Sex (at birth) |  |  |  |  |  |  |
| Men | 2,018 | 219 | 10.9 | 1,799 | 89.1 | 0.015<sup>a</sup> |
| Women | 2,260 | 300 | 13.3 | 1,960 | 86.7 |  |
| Unknown | 6 | 0 | 0.0 | 6 | 100.0 |  |
| Age (years) |  |  |  |  |  | 0.681 |
| Mean (SD) | 4,284 | 60.6 | 13.8 | 62.0 | 13.7 |  |
| European region |  |  |  |  |  | <0.001 |
| Central and Eastern | 1,413 | 138 | 9.8 | 1,275 | 90.2 |  |
| Western | 1,006 | 157 | 15.6<sup>b</sup> | 849 | 84.4 |  |
| Northern | 865 | 128 | 14.8<sup>b</sup> | 737 | 85.2 |  |
| Southern | 1,000 | 96 | 9.6 | 904 | 90.4 |  |
| Living situation |  |  |  |  |  | 0.073<sup>a</sup> |
| Alone | 794 | 103 | 13.0 | 691 | 87.0 |  |
| With spouse/partner | 1,879 | 223 | 11.9 | 1,656 | 88.1 |  |
| With spouse/partner and child(ren) | 1,063 | 114 | 10.7 | 949 | 89.3 |  |
| With other adult(s) and/or child(ren) | 356 | 53 | 14.9 | 303 | 85.1 |  |
| Other | 132 | 23 | 17.4 | 109 | 82.6 |  |
| Unknown | 60 | 3 | 5.0 | 57 | 95.0 |  |
| Highest level of education completed |  |  |  |  |  | <0.001<sup>a</sup> |
| None or primary | 366 | 32 | 8.7 | 334 | 91.3 |  |
| Lower secondary | 704 | 69 | 9.8 | 635 | 90.2 |  |
| Upper secondary (e.g., High School) | 1,481 | 148 | 10.0 | 1,333 | 90.0 |  |
| Post-secondary (e.g., College/University/Master/PhD) | 1,662 | 265 | 15.9<sup>b</sup> | 1,397 | 84.1 |  |
| Unknown | 71 | 5 | 7.0 | 66 | 93.0 |  |
| CIE - Original primary cancer diagnosis |  |  |  |  |  | 0.002<sup>a</sup> |
| Thoracic cancer (C30–C39) | 546 | 48 | 8.8 | 498 | 91.2 |  |
| Breast cancer (C50) | 869 | 134 | 15.4<sup>b</sup> | 735 | 84.6 |  |
| Digestive cancer (C15–C26) | 935 | 100 | 10.7 | 835 | 89.3 |  |
| Hematological cancer (C81–C96) | 451 | 51 | 11.3 | 400 | 88.7 |  |
| Genito-urinary cancer (C51–C68) | 972 | 114 | 11.7 | 858 | 88.3 |  |
| Other cancer (all other C codes, D00–D48) | 504 | 72 | 14.3 | 432 | 85.7 |  |
| Unknown | 7 | 0 | 0.0 | 7 | 100.0 |  |

Abbreviations: SD standard deviation, PhD Doctor of Philosophy, CIE Clinical Information Element
<sup>a</sup>Chi-squared *p*-value calculated excluding the ‘Unknown; skipped by the patient or missing value’ category.
<sup>b</sup>Post-hoc analyses using unadjusted standardized residuals; \|z\|> 1.96 indicate cells that contribute significantly to the overall group difference.

### WISP translation and coding

A total of 954 WISP responses were translated into English, of which six (0.6%) required further revision following review by native-speaking reviewers at the recruitment sites. For coding, the second researcher (HH) agreed with the initial code assigned by the first researcher (LRC) for 920 (96.4%) of the 954 WISP responses. The remaining 34 (3.6%) responses were discussed by the researchers, of which 14 (1.5% of all responses) were subsequently recoded by consensus within the research team. Three new symptom/problems categories not included in the WISP coding system were developed for this study: weight gain, hair loss, and workplace problems.

### Prevalence and severity of symptoms/problems reported on WISP

The 519 participants who completed the WISP provided a total of 954 symptoms/problems. As shown in Figure 1, 43.1% (411/954) concerned symptoms/problems already covered by existing items within the EUonQoL-Kit, 40.1% (383/954) were issues not covered (i.e., were additional) and 16.8% (160/954) were responses that could not be coded as symptoms/problems. These 160 responses were excluded from further analyses.

**Figure 1.**
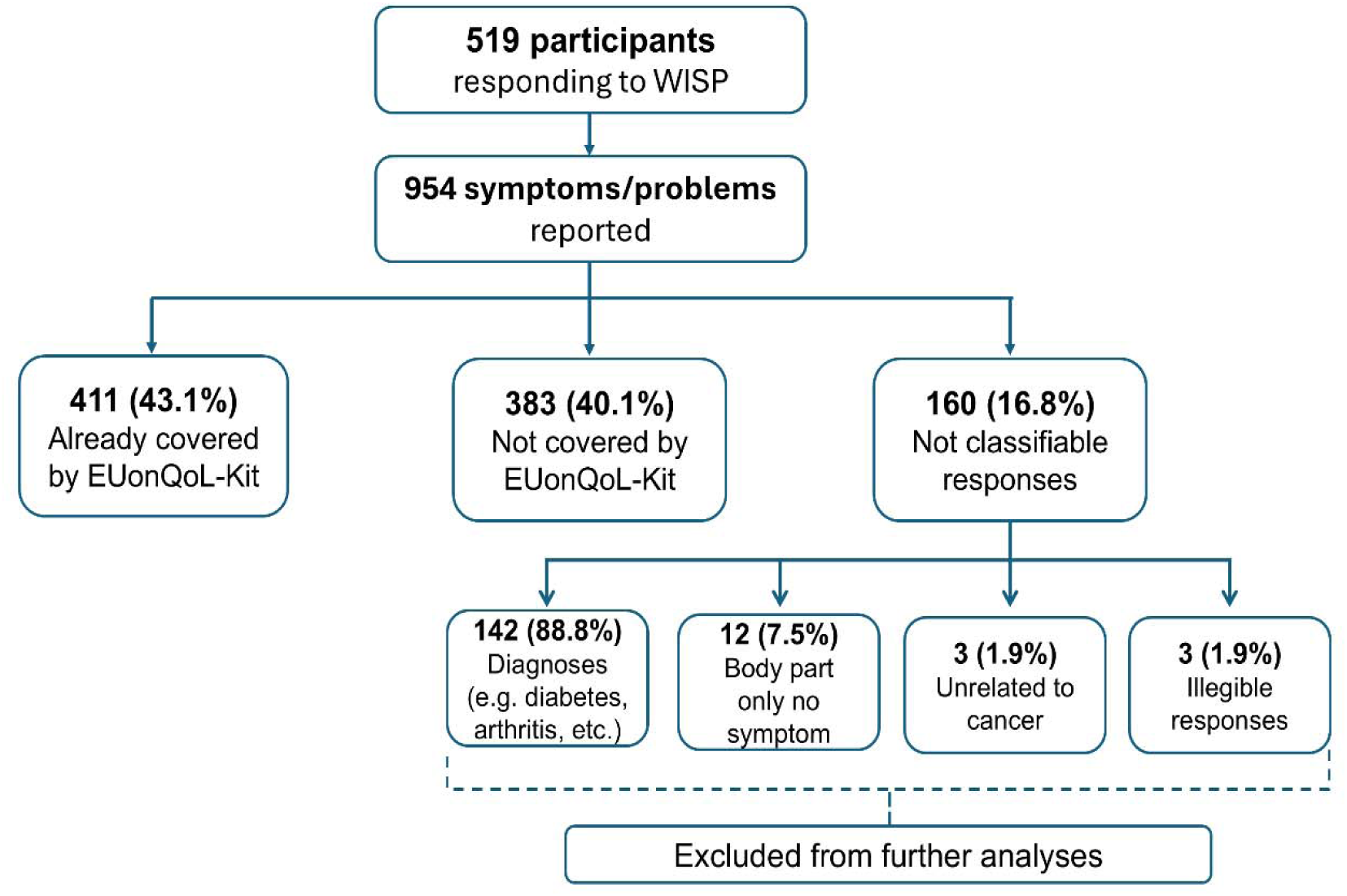
Classification of WISP responses listed by 519 participants among the 4,284 answering the EUonQoL-Kit v2.

The mean number of symptoms/problems reported on the WISP was 0.2 (median=0, range=0–6). The 519 participants who completed the WISP reported an average of 1.9 symptoms/problems on the WISP (median=2, range 1–6). Patients with cancer in active treatment or palliative care listed on average 1.9 symptoms/problems on WISP (median=2, range 1–6), whereas survivors listed on average 1.7 symptoms/problems (median=2, range 1–3).

Table 3 presents the prevalence of 58 symptom/problem categories identified from 794 symptom/problem reports, including symptoms/problems already covered or not covered by the EUonQoL-Kit v2. Prevalence was calculated as the proportion of the entire study population (N=4,284) reporting each symptom/problem category. The most prevalent additional symptoms/problems reported on the WISP were numbness/tingling (0.9%, n=38 out of entire sample of study participants), skin problems (0.7%, n=31) and dizziness (0.5%, n=20). The most prevalent symptoms/problems already covered by the EUonQoL-Kit v2 were pain (3.0%, n=130), impaired physical function (1.3%, n=54) and impaired emotional function (1.2%, n=52). The 54 issues classified as physical function comprised reduced mobility (33.3%), muscular weakness (20.4%), walking problems (18.5%), limitations in daily activities (16.7%), and balance/coordination problems (11.1%). Issues coded as impaired emotional function (n=52) comprise feeling sad (25.0%), anxious (23.1%), stressed (17.3%), unspecified emotional issue (15.4%), fearful (9.6%) and irritated (9.6 %).

**Table 3.** Prevalence of 58 symptom/problem categories reported on the EORTC WISP among participants in the EUonQoL-Kit validation study, overall and according to target population. A total of 794 symptoms/problems were reported by 519 of the 4,284 participants.

| 58 symptom/problem categories | Prevalence in 4,284 participants answering EUonQoL-Kit |  | Target populations |  |  |  |  |  |
|---|---|---|---|---|---|---|---|---|
|  |  |  | Active treatment (N=1,747) |  | Survivors (N=1,255) |  | Palliative care (N=1,282) |  |
|  | N | % | N | % | N | % | N | % |
| *Symptoms/problems not covered by the EUonQoL-Kit* |  |  |  |  |  |  |  |  |
| Numbness/tingling | 38 | 0.9 | 21 | 1.2 | 8 | 0.6 | 9 | 0.7 |
| Skin problems | 31 | 0.7 | 19 | 1.1 | 5 | 0.4 | 7 | 0.5 |
| Dizziness | 20 | 0.5 | 9 | 0.5 | 6 | 0.5 | 5 | 0.4 |
| Indigestion | 18 | 0.4 | 4 | 0.2 | 8 | 0.6 | 6 | 0.5 |
| Edema | 16 | 0.4 | 6 | 0.3 | 2 | 0.2 | 8 | 0.6 |
| Cough | 16 | 0.4 | 7 | 0.4 | 3 | 0.2 | 6 | 0.5 |
| Incontinence<sup>a</sup> | 16 | 0.4 | 4 | 0.2 | 9 | 0.7 | 3 | 0.2 |
| Vision problems | 15 | 0.4 | 4 | 0.2 | 6 | 0.5 | 5 | 0.4 |
| Bleeding | 15 | 0.4 | 8 | 0.5 | 2 | 0.2 | 5 | 0.4 |
| Itching | 14 | 0.3 | 5 | 0.3 | 2 | 0.2 | 7 | 0.5 |
| Urinary problems | 14 | 0.3 | 5 | 0.3 | 5 | 0.4 | 4 | 0.3 |
| Headache | 13 | 0.3 | 3 | 0.2 | 8 | 0.6 | 2 | 0.2 |
| Dysphagia | 12 | 0.3 | 4 | 0.2 | 3 | 0.2 | 5 | 0.4 |
| Existential problems | 10 | 0.2 | 4 | 0.2 | 1 | 0.1 | 5 | 0.4 |
| Dry mouth | 10 | 0.2 | 5 | 0.3 | 4 | 0.3 | 1 | 0.1 |
| Weight gain | 10 | 0.2 | 5 | 0.3 | 4 | 0.3 | 1 | 0.1 |
| Myoclonus<sup>b</sup> | 10 | 0.2 | 3 | 0.2 | 5 | 0.4 | 2 | 0.2 |
| Cognitive dysfunction | 9 | 0.2 | 4 | 0.2 | 2 | 0.2 | 3 | 0.2 |
| Fever | 8 | 0.2 | 4 | 0.2 | 2 | 0.2 | 2 | 0.2 |
| Reduced memory | 8 | 0.2 | 4 | 0.2 | 3 | 0.2 | 1 | 0.1 |
| Bloating | 7 | 0.2 | 1 | 0.1 | 1 | 0.1 | 5 | 0.4 |
| Sweats | 7 | 0.2 | 2 | 0.1 | 4 | 0.3 | 1 | 0.1 |
| Hair loss | 6 | 0.1 | 2 | 0.1 | 3 | 0.2 | 1 | 0.1 |
| Heartburn | 6 | 0,1 | 2 | 0.1 | 2 | 0.2 | 2 | 0.2 |
| Other eye symptoms | 6 | 0.1 | 1 | 0.1 | 3 | 0.2 | 2 | 0.2 |
| Hearing problems | 6 | 0.1 | 0 | 0.0 | 2 | 0.2 | 4 | 0.3 |
| Taste change | 4 | 0.1 | 2 | 0.1 | 1 | 0.1 | 1 | 0.1 |
| Other ear problems | 4 | 0.1 | 2 | 0.1 | 2 | 0.2 | 0 | 0.0 |
| Weight loss | 4 | 0.1 | 2 | 0.1 | 1 | 0.1 | 1 | 0.1 |
| Burning sensation | 4 | 0.1 | 2 | 0.1 | 1 | 0.1 | 1 | 0.1 |
| Confusion | 3 | 0.1 | 1 | 0.1 | 0 | 0.0 | 2 | 0.2 |
| Speaking problems | 3 | 0.1 | 1 | 0.1 | 1 | 0.1 | 1 | 0.1 |
| Shakiness | 3 | 0.1 | 1 | 0.1 | 2 | 0.2 | 0 | 0.0 |
| Restlessness | 2 | 0.0 | 0 | 0.0 | 0 | 0.0 | 2 | 0.2 |
| Sore mouth | 2 | 0.0 | 1 | 0.1 | 0 | 0.0 | 1 | 0.1 |
| Chills | 2 | 0.0 | 0 | 0.0 | 1 | 0.1 | 1 | 0.1 |
| Palpitations | 2 | 0.0 | 1 | 0.1 | 0 | 0.0 | 1 | 0.1 |
| Thirst | 1 | 0.0 | 1 | 0.1 | 0 | 0.0 | 0 | 0.0 |
| Burping | 1 | 0.0 | 0 | 0.0 | 1 | 0.1 | 0 | 0.0 |
| Hoarseness | 1 | 0.0 | 0 | 0.0 | 1 | 0.1 | 0 | 0.0 |
| Heaviness | 1 | 0.0 | 1 | 0.1 | 0 | 0.0 | 0 | 0.0 |
| *Symptoms/problems already covered by the EUonQoL-Kit* |  |  |  |  |  |  |  |  |
| Pain | 130 | 3.0 | 45 | 2.6 | 55 | 4.4 | 30 | 2.3 |
| Impaired physical function<sup>c</sup> | 54 | 1.3 | 14 | 0.8 | 18 | 1.4 | 22 | 1.7 |
| Impaired emotional function<sup>d</sup> | 52 | 1.2 | 16 | 0.9 | 23 | 1.8 | 13 | 1.0 |
| Fatigue | 41 | 1.0 | 20 | 1.1 | 8 | 0.6 | 13 | 1.0 |
| Sleeping difficulties | 21 | 0.5 | 9 | 0.5 | 7 | 0.6 | 5 | 0.4 |
| Dyspnea | 21 | 0.5 | 4 | 0.2 | 6 | 0.5 | 11 | 0.9 |
| Low satisfaction with care | 20 | 0.5 | 7 | 0.4 | 6 | 0.5 | 7 | 0.5 |
| Social problems | 13 | 0.3 | 7 | 0.4 | 3 | 0.2 | 3 | 0.2 |
| Diarrhea | 11 | 0.3 | 4 | 0.2 | 4 | 0.3 | 3 | 0.2 |
| Nausea | 10 | 0.2 | 6 | 0.3 | 1 | 0.1 | 3 | 0.2 |
| Vomiting | 9 | 0.2 | 2 | 0.1 | 2 | 0.2 | 5 | 0.4 |
| Constipation | 9 | 0.2 | 6 | 0.3 | 1 | 0.1 | 2 | 0.2 |
| Economic problems | 8 | 0.2 | 3 | 0.2 | 4 | 0.3 | 1 | 0.1 |
| Workplace problems | 5 | 0.1 | 0 | 0.0 | 4 | 0.3 | 1 | 0.1 |
| Lack of appetite | 4 | 0.1 | 1 | 0.1 | 1 | 0.1 | 2 | 0.2 |
| Concentration problems | 4 | 0.1 | 0 | 0.0 | 1 | 0.1 | 3 | 0.2 |
| Sexual problems | 4 | 0.1 | 0 | 0.0 | 4 | 0.3 | 0 | 0.0 |
| Total | 794 | 100 | 295 | 37.2 | 262 | 33.0 | 237 | 29.8 |

N represents the number of unique participants reporting each symptom/problem category; 519 WISP respondents generated 794 symptom/problem reports classified into 58 categories; percentages use the relevant total population as the denominator
<sup>a</sup>Including urinary, stool and unspecified incontinence
<sup>b</sup>Including muscle cramps and spasms
<sup>c</sup>Including balance/coordination problems, muscular weakness, reduced mobility and walking problems
<sup>d</sup>Including feeling sad, anxious, stressed, fearful, irritated and unspecified emotional issue

Overall, most of the symptoms/problems were rated as moderate (51.6%, n=410) or severe (30.5%, n=242) and a small proportion as mild (17.9%, n=142). Further details on the severity of the 58 symptom/problem categories are presented in Table 4.

**Table 4.** Severity of 58 symptom/problem categories reported on the EORTC WISP among participants in the EUonQoL-Kit validation study. A total of 794 symptoms/problems were reported by 519 of the 4,284 participants.

| 58 symptom/problems categories | Severity |  |  |  |  |  |  |
|---|---|---|---|---|---|---|---|
|  |  | Mild |  | Moderate |  | Severe |  |
|  | Total | N | % | N | % | N | % |
| *Symptoms/problems not covered by the EUonQoL-Kit* |  |  |  |  |  |  |  |
| Numbness/tingling | 38 | 8 | 21.1 | 19 | 50.0 | 11 | 28.9 |
| Skin problems | 31 | 3 | 9.7 | 19 | 61.3 | 9 | 29.0 |
| Dizziness | 20 | 6 | 30.0 | 10 | 50.0 | 4 | 20.0 |
| Indigestion | 18 | 0 | 0.0 | 10 | 55.6 | 8 | 44.4 |
| Edema | 16 | 1 | 6.3 | 10 | 62.5 | 5 | 31.3 |
| Cough | 16 | 4 | 25.0 | 9 | 56.3 | 3 | 18.8 |
| Incontinence<sup>a</sup> | 16 | 1 | 6.3 | 8 | 50.0 | 7 | 43.8 |
| Vision problems | 15 | 2 | 13.3 | 7 | 46.7 | 6 | 40.0 |
| Bleeding | 15 | 7 | 46.7 | 6 | 40.0 | 2 | 13.3 |
| Itching | 14 | 2 | 14.3 | 9 | 64.3 | 3 | 21.4 |
| Urinary problems | 14 | 2 | 14.3 | 6 | 42.9 | 6 | 42.9 |
| Headache | 13 | 3 | 23.1 | 8 | 61.5 | 2 | 15.4 |
| Dysphagia | 12 | 2 | 16.7 | 6 | 50.0 | 4 | 33.3 |
| Existential problems | 10 | 1 | 10.0 | 2 | 20.0 | 7 | 70.0 |
| Dry mouth | 10 | 3 | 30.0 | 4 | 40.0 | 3 | 30.0 |
| Weight gain | 10 | 2 | 20.0 | 5 | 50.0 | 3 | 30.0 |
| Myoclonus<sup>b</sup> | 10 | 0 | 0.0 | 5 | 50.0 | 5 | 50.0 |
| Cognitive dysfunction | 9 | 1 | 11.1 | 2 | 22.2 | 6 | 66.7 |
| Fever | 8 | 5 | 62.5 | 1 | 12.5 | 2 | 25.0 |
| Reduced memory | 8 | 1 | 12.5 | 7 | 87.5 | 0 | 0.0 |
| Bloating | 7 | 0 | 0.0 | 6 | 85.7 | 1 | 14.3 |
| Sweats | 7 | 2 | 28.6 | 4 | 57.1 | 1 | 14.3 |
| Hair loss | 6 | 2 | 33.3 | 4 | 66.7 | 0 | 0.0 |
| Heartburn | 6 | 2 | 33.3 | 4 | 66.7 | 0 | 0.0 |
| Other eye symptoms | 6 | 1 | 16.7 | 5 | 83.3 | 0 | 0.0 |
| Hearing problems | 6 | 1 | 16.7 | 3 | 50.0 | 2 | 33.3 |
| Taste change | 4 | 1 | 25.0 | 1 | 25.0 | 2 | 50.0 |
| Other ear problems | 4 | 2 | 50.0 | 2 | 50.0 | 0 | 0.0 |
| Weight loss | 4 | 0 | 0.0 | 3 | 75.0 | 1 | 25.0 |
| Burning sensation | 4 | 0 | 0.0 | 4 | 100 | 0 | 0.0 |
| Confusion | 3 | 0 | 0.0 | 3 | 100 | 0 | 0.0 |
| Speaking problems | 3 | 0 | 0.0 | 3 | 100 | 0 | 0.0 |
| Shakiness | 3 | 2 | 66.7 | 1 | 33.3 | 0 | 0.0 |
| Restlessness | 2 | 0 | 0.0 | 2 | 100 | 0 | 0.0 |
| Sore mouth | 2 | 0 | 0.0 | 2 | 100 | 0 | 0.0 |
| Chills | 2 | 0 | 0.0 | 0 | 0.0 | 2 | 100 |
| Palpitations | 2 | 1 | 50.0 | 1 | 50.0 | 0 | 0.0 |
| Thirst | 1 | 0 | 0.0 | 1 | 100 | 0 | 0.0 |
| Burping | 1 | 1 | 100 | 0 | 0.0 | 0 | 0.0 |
| Hoarseness | 1 | 0 | 0.0 | 1 | 100 | 0 | 0.0 |
| Heaviness | 1 | 1 | 100 | 0 | 0.0 | 0 | 0.0 |
| *Symptoms/problems already covered by the EUonQoL-Kit* |  |  |  |  |  |  |  |
| Pain | 130 | 27 | 20.8 | 73 | 56.2 | 30 | 23.1 |
| Impaired physical function<sup>c</sup> | 54 | 9 | 16.7 | 21 | 38.9 | 24 | 44.4 |
| Impaired emotional function<sup>d</sup> | 52 | 9 | 17.3 | 27 | 51.9 | 16 | 30.8 |
| Fatigue | 41 | 6 | 14.6 | 27 | 65.9 | 8 | 19.5 |
| Sleeping difficulties | 21 | 5 | 23.8 | 11 | 52.4 | 5 | 23.8 |
| Dyspnea | 21 | 8 | 38.1 | 8 | 38.1 | 5 | 23.8 |
| Low satisfaction with care | 20 | 1 | 5.0 | 6 | 30.0 | 13 | 65.0 |
| Social problems | 13 | 1 | 7.7 | 5 | 38.5 | 7 | 53.8 |
| Diarrhea | 11 | 3 | 27.3 | 5 | 45.5 | 3 | 27.3 |
| Nausea | 10 | 3 | 30.0 | 4 | 40.0 | 3 | 30.0 |
| Vomiting | 9 | 0 | 0.0 | 4 | 44.4 | 5 | 55.6 |
| Constipation | 9 | 0 | 0.0 | 8 | 88.9 | 1 | 11.1 |
| Economic problems | 8 | 0 | 0.0 | 3 | 37.5 | 5 | 62.5 |
| Workplace problems | 5 | 0 | 0.0 | 1 | 20.0 | 4 | 80.0 |
| Lack of appetite | 4 | 0 | 0.0 | 2 | 50.0 | 2 | 50.0 |
| Concentration problems | 4 | 0 | 0.0 | 2 | 50.0 | 2 | 50.0 |
| Sexual problems | 4 | 0 | 0.0 | 0 | 0.0 | 4 | 100 |
| Total | 794 | 142 | 17.9 | 410 | 51.6 | 242 | 30.5 |

N represents the number of unique participants reporting each symptom/problem category; 519 WISP respondents generated 794 symptom/problem reports classified into 58 categories
<sup>a</sup>Including urinary, stool and unspecified incontinence
<sup>b</sup>Including muscle cramps and spasms
<sup>c</sup>Including balance/coordination problems, muscular weakness, reduced mobility and walking problems
<sup>d</sup>Including feeling sad, anxious, stressed, fearful, irritated and unspecified emotional issue

### Comparison of symptom prevalence

The prevalence of the ten most common additional symptoms/problems reported on the WISP instrument is presented in Table 3. Some descriptive variation was observed in the prevalence of these frequent additional symptoms/problems across target populations. Numbness/tingling, skin problems and bleeding had the highest observed prevalence among patients receiving active treatment (1.2%, 1.1% and 0.5% respectively), while dizziness had the same observed prevalence among patients receiving active treatment and survivors (0.5% in both groups). Indigestion (0.6%), incontinence (0.7%), and vision problems (0.5%) had the highest observed prevalence among survivors, whereas edema (0.6%), cough (0.5%), and itching (0.5%) had the highest observed prevalence among patients receiving palliative care.

Exploratory comparisons using Fisher’s exact tests showed no statistically significant differences in the prevalence of any of the ten most reported additional symptoms/problems across the three target populations with *p* values that ranged from 0.130 for edema to 0.960 for dizziness.

## Discussion

### Main findings

This descriptive study investigated symptoms/problems reported through the EORTC WISP instrument in addition to those captured by the EUonQoL-Kit v2, assessed differences in background characteristics between participants who did and did not report on the WISP and compared the reporting of additional symptom/problem across three target populations.

The decision to include the WISP in the EUonQoL-Kit v2 was in response to two concerns; one to ensure the coverage of the instrument was succinct and relevant to all cancer populations whilst allowing for additional issues to be reported on and secondly, enabled the opportunity to establish if there were any missing issues which should been included in future versions. Overall, the WISP identified 383 symptoms/problems not covered by the EUonQoL-kit v2, accounting for 40.1% of the 954 symptoms/problems reported on WISP during the validation study. However, these uncovered symptoms/problems were reported by relatively few participants with the most common issue, numbness/tingling, being reported by <1% of the study population. Interestingly, pain, fatigue, and impact on physical and emotional function, although already comprehensively covered by the EUonQoL-Kit v2, were the most identified issues using WISP. This highlights their substantial burden in cancer populations and is consistent with previous WISP studies, which similarly identified these as prevalent issues among cancer patients in active or palliative treatment [16–18]. These findings illustrate the usefulness of the WISP instrument as a complement to standardized HRQoL assessment and support the initial issue-selection criteria used in the development of the EUonQoL-Kit v2, indicating that no major highly prevalent symptoms or concerns appear to have been missed.

Despite the WISP completion rate was lower in this study (12% of 4,284 participants) than in previous studies using the WISP alongside the 15-item EORTC QLQ-C15-PAL questionnaire, (33– 36%) [17, 18], likely reflecting the greater length of the EUonQoL-Kit v2. However, among WISP respondents, the mean number of symptoms/problems reported was comparable to previous research (1.9 among 519 participants reporting at least one symptom/problem vs. 1.6 among 1,788 patients receiving palliative cancer care) [17]. Thus, the average number of symptoms/problems reported among WISP respondents were consistent with previous findings, even though respondent fatigue or other factors may have contributed to underreporting symptoms/problems during the validation study.

Women, participants from Western and Northern Europe, highly educated and those diagnosed with breast cancer were more likely to respond to the WISP than other subgroups. These differences suggest that engagement with an open-ended question was not uniform across the study population. Previous oncology studies have similarly shown that women and patients with breast cancer are more likely to report emotional, social and financial concerns, as well as issues related to cancer care, when given the opportunities for free-text reporting [22, 23]. In contract, previous WISP research found no overall association between reporting any additional symptom/problem and sex, age, or cancer diagnosis; however, certain types of cancer were associated with specific symptoms/problems [24]. Because WISP relies on spontaneous symptom reporting, differences in the reporting may reflect not only differences in symptom burden but also how patients perceive the relevance of their symptoms, their ability to recall symptoms at the time of completion, and their cultural or linguistic comfort in expressing health concerns in an open-text format, cognitive factors that should be consider when interpreting heterogeneity among WISP respondents in future research.

Although Fisher’s exact tests did not identify statistically significant differences in the prevalence of the ten most common additional symptoms/problems across the three target populations, the descriptive patterns provides context reported in different care settings. Patients in active treatment frequently reported numbness/tingling, skin problems and bleeding. While a previous WISP validation study found numbness/tingling to be more common among patient with cancer receiving palliative care, skin problems and bleeding were consistently reported during active treatment [16]. These concerns are consistent with the range of symptoms and adverse effects that may accompany anticancer treatments, including targeted therapies [25, 26].

In the current study, cancer survivors frequently reported indigestion, incontinence, and vision problems on the WISP, consistent with literature identifying gastrointestinal, urinary, and visual problems as long-term sequelae of cancer treatment such as surgery, radiotherapy, chemotherapy and corticosteroid use [27]. An exploratory study of 3,300 cancer survivors similarly found that urinary incontinence was among the most common physical problems reported via open-ended question after completing a PRO questionnaire [28]. Patients in palliative care commonly reported edema, cough, and itching, a finding that aligns with previous studies identifying these symptoms among the most frequent issues reported through open-ended questions by patients with advanced cancer [17, 29, 30].

Notably, 82% of symptoms/problems reported through WISP were rated as moderate to severe, similar to the 78–85% reported in previous WISP studies [16–18]. This suggests that patients do not necessarily use an open-ended question to list every symptom they experience but may instead focus on problems they consider particularly important or burdensome.

### Strengths and limitations

A strength of this study is the evaluation of WISP beyond palliative care settings in a large, multinational European sample and its first evaluation in a digital format. This study also identified three additional symptom/problem categories not included in the existing EORTC WISP coding system: weight gain, hair loss, and workplace problems. Even though the EUonQoL-Kit v2 included an item assessing support received from employers, it seems this item not fully captured diverse workplace problems experienced by the participants, highlighting the value of open-ended measures in detecting concerns that may not be extensively covered by standardized questionnaires.

However, some limitations require consideration. First, relatively few participants reported individual symptoms/problems through WISP, limiting the statistical power for comparisons across the target populations. Thus, the specific findings on symptom prevalence and severity must be interpreted descriptively. The absence of statistically significant differences across the three target populations reporting the ten most frequent additional symptoms/problems should not be interpreted as evidence of equivalence of these groups.

Second, only 12.1% of participants reported at least one symptom/problem through WISP, and WISP respondents differed from nonrespondents in several background characteristics. The additional identified symptoms/problems may therefore reflect, in part, differences in willingness or ability to respond to an open-ended question rather than differences in symptom burden alone.

Finally, the digital interface on WISP administration influenced participant behavior, as 41 of 519 participants (7.9%) entered multiple symptoms/problems in the initial response field, including 9 participants who reported more than the three requested symptoms/problems. This may indicate that some participants did not recognize the option to report additional symptoms/problems in subsequent steps. This user experience informed subsequent development of the EUonQoL-Kit V3, in which the WISP was retained because of its ability to capture symptoms/problems not otherwise covered by the toolkit, while changes to the user interface and instructions were introduced to improve usability.

## Conclusions

The EORTC WISP complemented standardized PRO assessment by capturing additional symptoms/problems not covered by the closed-ended items of the EUonQoL-Kit v2. Despite the relatively small proportion of participants reporting additional symptoms/problems, the findings highlight the value of open-ended reporting in identifying issues that may be overlooked by standardized HRQoL measures.

Descriptive variation in reported symptoms/problems across patients in active treatment, survivors, and patients in palliative care further suggests that WISP provides a flexible way to elicit population-specific concerns without requiring additional predefined items for all patients. However, no statistically significant differences were observed in the prevalence of commonly reported additional symptoms/problems between target populations. Integrating WISP into the final toolkit may therefore enhance the comprehensiveness and patient-centeredness of HRQoL assessment across diverse European cancer populations while preserving the efficiency of standardized questionnaires.

## Declarations

### Author contributions

Giovanni Apolone, Cinzia Brunelli, Sara Iadecola and Mogens Groenvold contributed to study conceptualization and design. Anne Brédart, Federico Borgogni, Ana Carneiro, Tudor Ciuleanu, Michal Chovanec, Philip R. Debruyne, Daniele Doege, Theres Fey, Orsolya Horváth, Angelos Kassianos, Tomáš Kazda, Ekaterina Kldiashvili, Sandra Lejniece, Iwona Ługowska, Johanna Mattson, Milana M. Asković, Helidon Nina, Helle Pappot, Ivaylo Petrov, Vadim Pogonet, Ivica Ratoša, Alessandro Rizzo, Marie Robert, Gerald Sendlhofer, Antonio Tanzilli, Vahur Valvere, Ines Vaz-Luis contributed to investigation. Leslye Rojas-Concha and Hayat Hamzeh contributed to data curation and formal analysis. Cinzia Brunelli, Augusto Caraceni, Montserrat Ferrer, Mogens Groenvold, Marianne G. Guren, Gabriella Pravettoni and Hugo Vachon as members of the Executive Commitee contributed to project administration, funding acquisition and resources. Alexandra Gilbert and Mogens Groenvold contributed to supervision. Leslye Rojas-Concha contributed to writing – original draft in close collaboration with Mathilde Adsersen, Maiken B. Hansen and Morten A. Petersen. Carina Dantas, Luana Caselli, Olatz Garin, Chiara Marzorati, Inke Minnée-van Braak and Emma Nicklin contributed to writing – review and editing the manuscript. All authors commented on previous versions and approved the final manuscript.

## Acknowledgments

The authors would like to acknowledge the following members of the EUonQoL Working Group: Olga Balaoura; Sarah Ball; Prof. Maria José Bento; Prof. Christian Brandts; Gennaro Ciliberto; Norbert Couespel; Dr. Claire Donohoe; Audrius Dulskas; Dr. Nicole Erickson; Laura Gangeri; Dr. Mahmut Gumus; Iris van der Heide; Caitriona Higgins; Stein Kaasa; Tapani Kalmi; Prof. Maria Litwiniuk; Claudio Lombardo; Aline Machiavelli; Elise Martin; Andrea Pace; Liina Pääbo; Madeline Pe; Ricardo Pietrobon; Laura Pinnavaia; Prof. Lonneke van de Poll; Dr. Katarzyna Pogoda; Dr. Vesna Ramljak; Femke Van Schelven; Dr. Anna Sikora-Koperska; Aude Sirven; Prof. Laurentiu Simion; Eda Tanrıkulu Simsek; Melissa Thong; and Galina Velikova.

## Ethics approval and consent to participate

The multicenter validation study was conducted in accordance with the principles of the Declaration of Helsinki. The study protocol was approved by the Ethics Committee of the coordinating center, Fondazione IRCCS Istituto Nazionale dei Tumori di Milano (Comitato Etico Territoriale Lombardia 4; approval number INT 106/23). In addition, all 45 participating cancer centers obtained approval from their respective local ethics committees. The corresponding approval numbers are reported in the study protocol (Additional File 2; Apolone et al. [15]). A detailed list of the participating centers is available on ClinicalTrials.gov (study registration number: NCT05947903). Written informed consent was obtained from all participants enrolled in the validation study.

## Data availability

The data are not currently publicly available. In accordance with the study protocol, they will be made available once fully anonymized, five years after completion of data collection.

## References

1. Balitsky AK, Rayner D, Britto J, Lionel AC, Ginsberg L, Cho W, et al. Patient-Reported Outcome Measures in Cancer Care: An Updated Systematic Review and Meta-Analysis. JAMA Netw Open. 2024;7(8):e2424793. doi: 10.1001/jamanetworkopen.2024.24793.

2. Lehmann J, Dragan T, Rammant E, de Ligt KM, Lai-Kwon J, Lidington E, et al. Exploring the integration of patient-reported outcome measures in clinical practice: A cross-sectional survey of EORTC healthcare professionals. Eur J Cancer. 2025;220:115333. doi: 10.1016/j.ejca.2025.115333.

3. Graupner C, Kimman ML, Mul S, Slok AHM, Claessens D, Kleijnen J, et al. Patient outcomes, patient experiences and process indicators associated with the routine use of patient-reported outcome measures (PROMs) in cancer care: a systematic review. Support Care Cancer. 2021;29(2):573–93. doi: 10.1007/s00520-020-05695-4.

4. Gilbert A, Sebag-Montefiore D, Davidson S, Velikova G. Use of patient-reported outcomes to measure symptoms and health related quality of life in the clinic. Gynecol Oncol. 2015;136(3):429–39. doi: 10.1016/j.ygyno.2014.11.071.

5. Al Sayah F, Lahtinen M, Bonsel GJ, Ohinmaa A, Johnson JA. A multi-level approach for the use of routinely collected patient-reported outcome measures (PROMs) data in healthcare systems. J Patient Rep Outcomes. 2021;5(Suppl 2):98. doi: 10.1186/s41687-021-00375-1.

6. Haverman L, Luijten MAJ, Blackford AL, Absolom K, Basch EM, van Rossum MAJ, et al. Truth and dare: patients dare to tell the truth when using PROMs in clinical practice. Qual Life Res. 2024;33(12):3299–307. doi: 10.1007/s11136-024-03772-3.

7. Apolone G, Brunelli C. Quality of Life in Oncology: Measuring What Matters for Cancer Patients and Survivors in Europe: The EUonQol Project. J Ambul Care Manage. 2023;46(2):139–42. doi: 10.1097/jac.0000000000000449.

8. Vachon H, Giljević KM, Piccinin C, Shkodra M, Petersen MA, van Braak IM, et al. Evaluation of the psychometric properties of patient-reported outcome measures for the assessment of health-related quality of life across the European cancer continuum: a systematic review applying COSMIN methodology. Health Qual Life Outcomes 2026. doi: 10.1186/s12955-026-02616-7.

9. Lizano-Barrantes C, Amat-Fernandez C, Garin O, Luer-Aguila R, Pardo Y, Rojas-Concha L, et al. Needs and health-related quality of life domains relevant to people in Europe with advanced cancer in need of palliative care: a systematic review of qualitative research. Qual Life Res. 2026;35(3):74. doi: 10.1007/s11136-025-04129-0.

10. Lizano-Barrantes C, Amat-Fernandez C, Garin O, Pardo Y, Périz-Colón Ó, Saint-Jean PN, et al. Health-related quality of life domains relevant to people in Europe undergoing cancer treatment: a systematic review of qualitative research. Qual Life Res. 2026;35(6):158. doi: 10.1007/s11136-026-04201-3.

11. Amat-Fernandez C, Garin O, Luer-Aguila R, Pardo Y, Briseño R, Lizano-Barrantes C, et al. Systematic review of the needs and health-related quality of life domains relevant to people surviving cancer in Europe. Qual Life Res. 2025;34(4):913–36. doi: 10.1007/s11136-024-03884-w.

12. Brédart A, Martin E, Gillanders E, Bedding C, Alfieri S, Apolone G, et al. Exploring health-related quality of life with patients and survivors across the cancer care continuum - the EUonQoL-Kit mixed-method interview study. Journal of Epidemiology and Population Health. 2026;74(5):203787. doi: 10.1016/j.jeph.2026.203787.

13. Sweegers MG, de Jongh E, Bedding C, Nicklin E, Doege D, Alfieri S, et al. Development of a unified system for assessing health related quality of life across the cancer care continuum: the EUonQoL Delphi study to identify priorities for quality of life domains. J Patient Rep Outcomes. 2025;9(1):70. doi: 10.1186/s41687-025-00907-z.

14. Nicklin E, Bedding C, Alfieri S, Apolone G, Arndt V, Bos N, et al. The development of the European Toolkit for the self-assessment of quality of life across the cancer care continuum (EUonQoL-Kit). Eur J Cancer. 2026. doi: 10.1016/j.ejca.2026.116677.

15. Apolone G, Costantini M, Caselli L, Bos N, Caraceni A, Ciliberto G, et al. Validation of the European Oncology toolkit for the self-assessment of Quality of Life (EUonQoL-Kit) in cancer patients and survivors: study protocol of a pan European survey. BMC Public Health. 2024;24(1):3517. doi: 10.1186/s12889-024-21008-4.

16. Rojas-Concha L, Arrarrás JI, Conroy T, Chalk T, Guberti M, Holzner B, et al. Acceptability and usefulness of the EORTC ‘Write In three Symptoms/Problems’ (WISP): a brief open-ended instrument for symptom assessment in cancer patients. Health Qual Life Outcomes 2024;22(1):28. doi: 10.1186/s12955-024-02244-z.

17. Rojas-Concha L, Hansen MB, Petersen MA, Groenvold M. Which symptoms and problems do advanced cancer patients admitted to specialized palliative care report in addition to those included in the EORTC QLQ-C15-PAL? A register-based national study. Support Care Cancer. 2020;28(4):1725–35. doi: 10.1007/s00520-019-04976-x.

18. Rojas-Concha L, Hansen MB, Petersen MA, Groenvold M. Symptoms of advanced cancer in palliative medicine: a longitudinal study. BMJ Support Palliat Care. 2023;13(e2):e415–e27. doi: 10.1136/bmjspcare-2021-002999.

19. Rojas-Concha L, Petersen MA, Bedding C, Velikova G, Gilbert A, Sweegers MG, et al. Integrating the EORTC Computerized Adaptive Testing (CAT) Core in the European Oncology Quality of Life toolkit (EUonQoL-Kit) for the assessment of health-related quality of life in cancer patients and survivors. PREPRINT (Version 1) available at Research Square 31 March 2026. doi: 10.21203/rs.3.rs-9211206/v1.

20. Linlin L. Artificial Intelligence Translator DeepL Translation Quality Control. Procedia Comput Sci. 2024;247:710–7. doi: 10.1016/j.procs.2024.10.086.

21. Rojas-Concha L, Gilbert A, Kuliś D, Piccini C, Groenvold M: EORTC Quality of Life Group Guideline for the use and interpretation of the EORTC WISP instrument https://qol.eortc.org/manual/eortc-wisp-guideline/ (2024). Accessed 22 April 2026.

22. Burg MA, Adorno G, Lopez EDS, Loerzel V, Stein K, Wallace C, et al. Current unmet needs of cancer survivors: Analysis of open-ended responses to the American Cancer Society Study of Cancer Survivors II. Cancer. 2015;121(4):623–30. doi: 10.1002/cncr.28951.

23. Cunningham M, Wells M. Qualitative analysis of 6961 free-text comments from the first National Cancer Patient Experience Survey in Scotland. BMJ Open. 2017;7(6):e015726. doi: 10.1136/bmjopen-2016-015726.

24. Rojas-Concha L, Petersen MA, Groenvold M. Characteristics of patients reporting symptoms by open-ended questions in specialist palliative care. Dan Med J. 2021;68(8):A12200916.

25. Tímár J, Uhlyarik A. On-target side effects of targeted therapeutics of cancer. Pathol Oncol Res. 2022;28:1610694. doi: 10.3389/pore.2022.1610694.

26. Yazbeck V, Alesi E, Myers J, Hackney MH, Cuttino L, Gewirtz DA. Chapter One - An overview of chemotoxicity and radiation toxicity in cancer therapy. Adv Cancer Res. 2022;155:1–27. doi: 10.1016/bs.acr.2022.03.007.

27. Miller KD, Triano LR. Medical Issues in Cancer Survivors—A Review. Cancer J. 2008;14(6):375–87. doi: 10.1097/PPO.0b013e31818ee3dc.

28. Corner J, Wagland R, Glaser A, Richards SM. Qualitative analysis of patients’ feedback from a PROMs survey of cancer patients in England. BMJ Open. 2013;3(4):e002316. doi: 10.1136/bmjopen-2012-002316.

29. Alsirafy SA, Abd El-Aal HH, Farag DE, Radwan RH, El-Sherief WA, Fawzy R. High symptom burden among patients with newly diagnosed incurable cancer in a developing country. J Pain Symptom Manage. 2016;51(5):e1–e5. doi: 10.1136/bmjspcare-2019-001880.

30. Homsi J, Walsh D, Rivera N, Rybicki LA, Nelson KA, Legrand SB, et al. Symptom evaluation in palliative medicine: patient report vs systematic assessment. Support Care Cancer. 2006;14(5):444–53. doi: 10.1007/s00520-005-0009-2.

